# Trends and Influences in women authorship of randomized controlled trials in rheumatology: a comprehensive analysis of all published RCTs from 2009 to 2023

**DOI:** 10.1101/2024.08.26.24312469

**Authors:** Kim Lauper, Diana Buitrago-Garcia, Delphine Courvoisier, Michele Iudici, Denis Mongin

## Abstract

**Objectives:** This study aims to examine the evolution and influencing factors of women’s authorship in randomized controlled trials (RCTs) published in rheumatology.

**Methods:** This study included all rheumatology RCTs published from 2009 to 2023. The gender of authors was determined using forenames and countries of affiliation via the gender API service. The percentage of women in RCT publications and its association with potential factors was assessed using generalized estimating equations, considering women gender as the main binary outcome and the RCT’s continent, international collaboration status, industrial funding, intervention type, sample size, journal adherence to ICMJE recommendations, impact factor, publication year, author’s non-academic affiliation, and author position as covariates.

**Results:** Among the 1,092 RCTs authored by 10,794 persons, women accounted for 34.1% of authors. Woman authorship was more frequent in African-based RCTs compared to North America, when the author had a non-academic affiliation and when the last author was a woman (1.83 [1.46, 2.29], +6.1 percentage points – pp). Woman authorship was less frequent in Asian and European-based RCTs, industry-funded RCTs (OR 0.64 [0.56-0.73]; -10.3pp). Women were less often in the last (0.63 [0.54-0.74]; -10.2 pp) and second to last author position (0.73 [0.62-0.85]; -7.3pp). There were no difference looking at international status or year of publication.

**Conclusion:** The overall presence of women authors was 34.1%. The stagnant year-over-year representation of women in RCTs, and the lower likelihood of a woman having a position as senior author, underscores the need for more effective strategies to bridge the gender gap. RCTs with a woman last author were more likely to have a woman first author, suggesting a potential role-model effect.

## Introduction

Gender disparities are a pervasive issue in the realm of academia, consistently observed across diverse scientific fields and affecting various facets of academic life. Women face diverse challenges such as achieving fewer tenured positions, higher attrition rates during their careers (1,2), limited access to research funding (3–5), reduced opportunities for publication (6) and less frequent participation in editorial boards of scientific journals (7). When they do publish, women tend to be underrepresented as last authors (8) and are less credited than men for their work (9).

In recent years, there has been some progress in narrowing the gender publication gap, but the pace of improvement has shown signs of slowing down (10), particularly exacerbated by the challenges posed by the COVID-19 pandemic (11,12). In medicine, numerous studies have documented similar trends across subfields (10,13,14) and in various medical subspecialties (15–21). The field of rheumatology is no exception (22). Women face a scarcity of invited speaking opportunities at major conferences (23), reduced likelihood of attaining professorial positions, and a diminished chance of securing research grants, particularly in the United States (24). While there has been a recent move towards gender equity among authors in medical publications, women continue to be underrepresented in national and international organizations (25,26), as last authors (22), and are notably less present in randomized controlled trials (RCTs) (27).

The underrepresentation of women in RCTs is concerning for multiple reasons. First, the gender gap in RCT participation can have repercussions for the quality of science itself. Previous studies suggest that minorities tend to produce more scientific novelties (28), and women can exhibit superior scientific conduct (29), emphasizing the importance of gender diversity in research. Second, at a more individual level, it can be detrimental to the academic careers of women, as participation in RCTs is considered a significant milestone, given their status as the gold standard for scientific evidence in medicine. Inclusion as a first or last author on an RCT can be pivotal for one’s academic trajectory.

In this work, we aim to evaluate the factors influencing women’s authorship using a comprehensive analysis of all RCTs published in rheumatology since 2009. The objectives of this study are to assess the association of women’s authorship with RCTs and journal-specific factors, analyze how woman authorship correlates with author characteristics, and examine the temporal trends in women’s authorship.

## Methods

### Identification of RCT and associated data

For this study we included all RCTs in rheumatology published from 2009 to 2023, based on a previous work concerned with RCT registration in rheumatology (30). Briefly, we identified RCTs using the highly sensitive search Strategy for identifying randomized trials in MEDLINE-Pubmed developed by the Cochrane Collaboration (31,32) within all journals referred in the category rheumatology of the 2022 *Journal Citation Reports* of Clarivate, or in combination with a list of rheumatologic conditions within the five top journals in internal medicine.

Author data information (name, surname, affiliation) was retrieved from Medline metadata. Also, we obtained the trial registration number (TRN), abstract and full text using a validated dictionary of regular expression patterns. Trial information for RCTs with a registration number was retrieved from the International Clinical Trials Registry Platform of the World Health Organization webpage (planned sample size, disease, countries involved, intervention, funding source, date of registration and, date of first enrollment). For non-registered RCTs, information was retrieved from published full text.

### Identification of gender of each author

Gender was assessed based on the first name and the country of affiliation of each author using the gender API service, demonstrated to be one of the most accurate tools available for this task (33,34). The gender prediction algorithm is based on a database of more than six million names within 190 countries. Each gender prediction is associated with a probability corresponding to the estimated gender frequency of the given first name in the specified country. When the first name was not reported, the gender was left missing. To account for the uncertainty of the gender determination, the analysis consisted of a pooled analysis of 50 datasets, where the gender was set to according to the probability given by the gender determination algorithm (*see statistical analysis section*).

### Outcome and covariates

The analysis is made at the author level and the main outcome in our study is whether a given author of a published RCT is a woman.

The covariates evaluated for their influence on women authorship in the main model were the geographical location of the RCT (Europe, Asia, North America, South America, Africa, Oceania, Transcontinental), the type of sponsorship (Industry sponsor or not), the type of intervention (pharmacological or not), planned sample size, compliance of the journal publishing the RCT with the International Committee of Medical Journal Editors recommendations for the conduct, reporting, editing and publication of scholarly work in medical journals (ICMJE recommendations), journal impact factor by year of publication categorized in three categories (between 0 and 2, between 2 and 5, or above 5), year of publication (expressed as years since 2009), if authors had a pharmaceutical industry affiliation, and author’s position in the author list (first, second, middle, penultimate, last). If the publication had only two authors, they were considered first and last; if the publication had three authors, first, middle, and last; if the publication had four authors, first, second, penultimate, and last. In cases with more than four authors, the first, second, penultimate, and last authors were classified accordingly, while all remaining authors were grouped as middle authors.

### Statistical analysis

The percentage of women that were authors in published RCTs and the effects of the considered covariates were determined using generalized estimating equations (GEE), with the main outcome as dependent variable and the covariates as independent variables. The publications were the clusters, and we considered an exchangeable correlation structure. A gaussian distribution was used to obtain an estimate corresponding to percentages for estimating differences in % (using a Gaussian link with robust SE)(35), and the models were also conducted using a binomial distribution and a logistic link function to obtain odds ratio. Both percentage and odds ratio are reported.

In a first model, the independent variables were: continent where the RCT was performed, journal adherence to the ICJME recommendations, journal impact factor, planned sample size, year of publication, type of sponsor, type of author affiliation (academic or non-academic), and author position. In a second model, the independent variables additionally included whether the last author was identified as a woman. To avoid spurious association between the outcome (authors being a woman) and this additional variable, all last authors were removed from the dataset for this model.

To account for the uncertainty in gender determination, 50 imputed datasets were built in which gender was imputed with the probability given by the gender determination algorithm of genderize API. If the first name was not provided or if the gender API did not provide gender with the associated probability, the gender was imputed using chained equation algorithm with predictive mean matching for numerical data and logistic regression imputation for categorical data, considering gender and all covariates in the model. Analysis was then performed on each imputed dataset, and estimates were pooled according to Rubin’s rule (36). All analysis were performed using R 4.3.1 (37), with the library *geepack* (38) for the generalized estimating equations and the library *mice* (39) for the multiple imputation. All data and analysis code are publicly available at the following Gitlab repository: https://gitlab.unige.ch/trial_integrity/gender_rct_public.

## Results

### Published RCTs

Of the 1,092 rheumatology RCTs published since 2009, 42.8% assessed a pharmacological intervention (Table 1). Concerning the geographic location of the RCT sites, 36.2% were based in Europe only, 24.1% in Asia, followed by 13.1% in North America and 11.5% were transcontinental RCTs. Only 1% of the included RCTs were based in Africa. The conditions most often evaluated were rheumatoid arthritis (20.3%), followed by osteoarthritis (20.1%) among others. There was an increase in the number of RCTs published across years.

**Table 1.** Characteristics of included randomized controlled trials (RCTs)

|  | Overall |
| --- | --- |
| Number of trials | 1092 |
| Intervention (%) |  |
| Drug | 467 (42.8) |
| Non Drug | 625 (57.2) |
| Sponsor (%) |  |
| Industry | 421 (38.6) |
| Non-industry | 631 (57.8) |
| Not reported | 40 (3.7) |
| Impact Factor |  |
| 0-2 | 114 (10.4) |
| 2-5 | 617 (56.6) |
| 5+ | 360 (33.0) |
| Geographic location (%) |  |
| Europe | 395 (36.2) |
| Asia | 263 (24.1) |
| North America | 143 (13.1) |
| Transcontinental | 126 (11.5) |
| Oceania | 81 (7.4) |
| South America | 43 (3.9) |
| Africa | 11 (1.0) |
| Not reported | 30 (2.7) |
| Medical condition (%) |  |
| Rheumatoid arthritis | 222 (20.3) |
| Osteoarthritis | 220 (20.1) |
| Fibromyalgia | 55 (5.0) |
| Lupus | 54 (4.9) |
| Low back pain | 46 (4.2) |
| Knee arthroplasty | 38 (3.5) |
| Psoriatic arthritis | 37 (3.4) |
| Systemic sclerosis | 27 (2.5) |
| Gout | 24 (2.2) |
| Spondyloarthritis | 24 (2.2) |
| Ankylosing spondylitis | 20 (1.8) |
| Fractures | 19 (1.7) |
| Multiple diseases | 19 (1.7) |
| Juvenile idiopathic arthritis | 18 (1.6) |
| Other type of arthritis | 15 (1.4) |
| Sjogren's syndrome | 15 (1.4) |
| Knee pain | 14 (1.3) |
| Anterior cruciate ligament Injuries | 12 (1.1) |
| Neck pain | 12 (1.1) |
| Subacromial impingement syndrome | 11 (1.0) |
| Other | 190 (17.4) |
| Year of publication (%) |  |
| 2009 | 14 (1.3) |
| 2010 | 32 (2.9) |
| 2011 | 45 (4.1) |
| 2012 | 49 (4.5) |
| 2013 | 57 (5.2) |
| 2014 | 66 (6.0) |
| 2015 | 67 (6.1) |
| 2016 | 81 (7.4) |
| 2017 | 98 (9.0) |
| 2018 | 71 (6.5) |
| 2019 | 103 (9.4) |
| 2020 | 97 (8.9) |
| 2021 | 159 (14.6) |
| 2022 | 153 (14.0) |

### Authorship

The included RCTs were authored by 10,794 individuals. Most of the authors were affiliated to a non-industry institution, and only 7 % of the authors were affiliated to an industry. Most authors were affiliated to institutions in the United States of America and Great Britain, followed by France, China, the Netherlands, Australia and Japan.

**Table 2.**
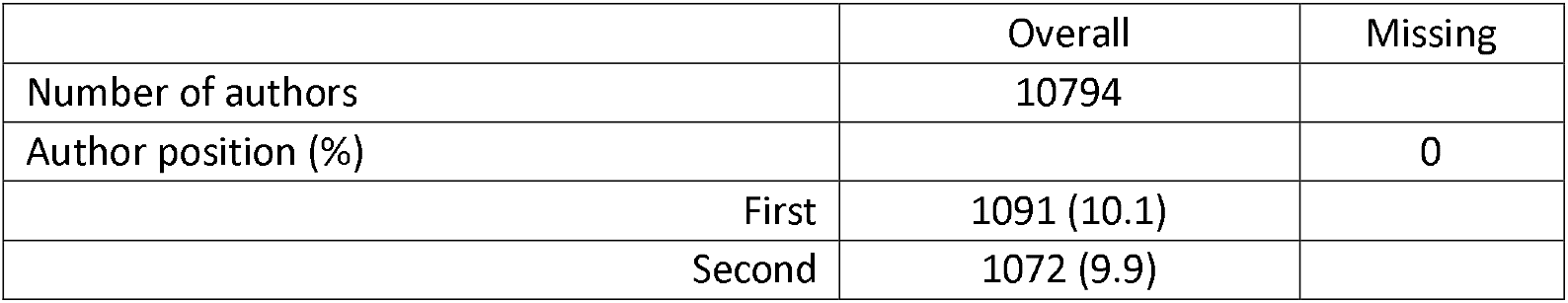
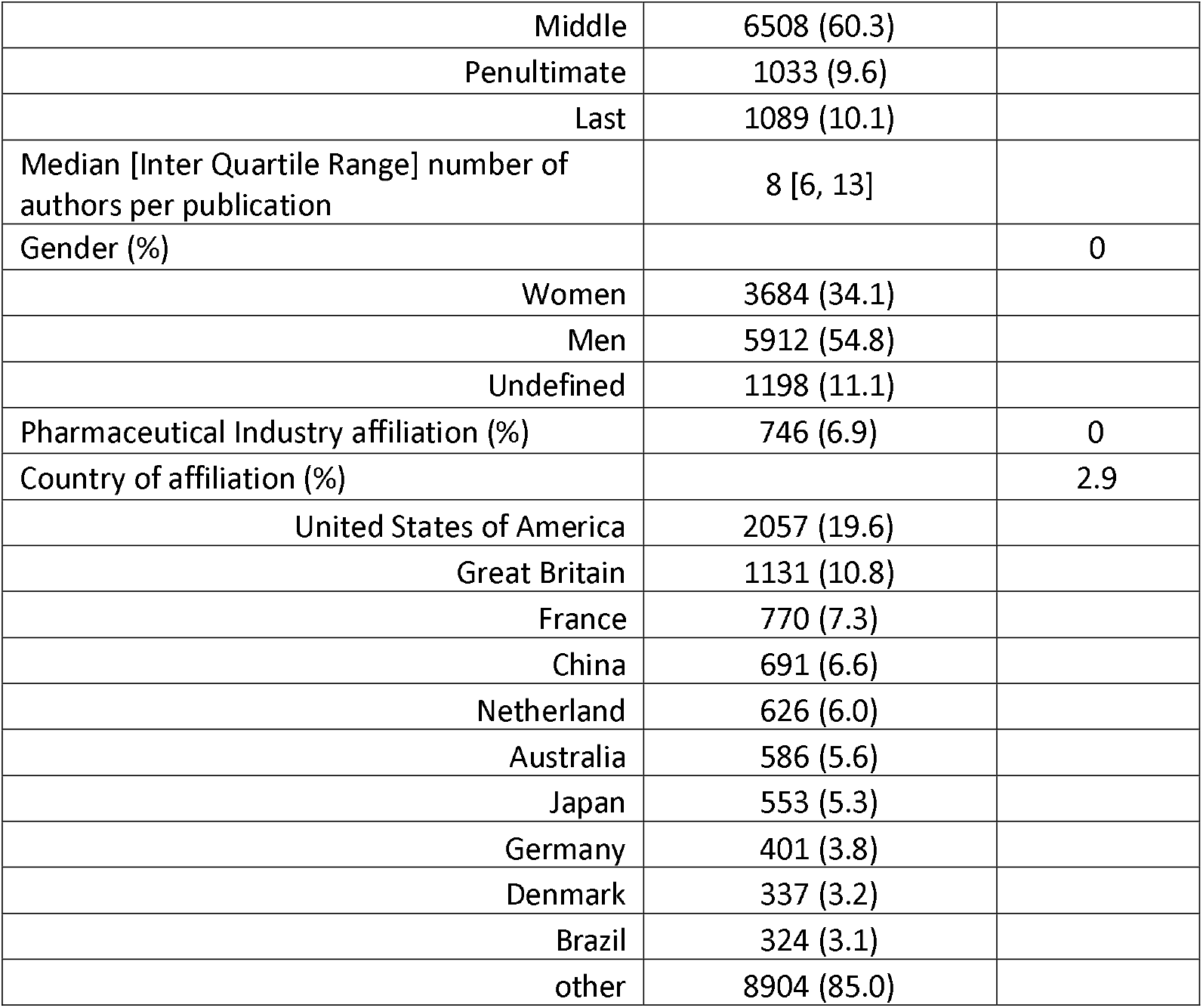
Description of the authors of the included RCTs

|  | Overall | Missing |
| --- | --- | --- |
| Number of authors | 10794 |  |
| Author position (%) |  | 0 |
| First | 1091 (10.1) |  |
| Second | 1072 (9.9) |  |
| Middle | 6508 (60.3) |  |
| Penultimate | 1033 (9.6) |  |
| Last | 1089 (10.1) |  |
| Median [Inter Quartile Range] number of authors per publication | 8 [6, 13] |  |
| Gender (%) |  | 0 |
| Women | 3684 (34.1) |  |
| Men | 5912 (54.8) |  |
| Undefined | 1198 (11.1) |  |
| Pharmaceutical Industry affiliation (%) | 746 (6.9) | 0 |
| Country of affiliation (%) |  | 2.9 |
| United States of America | 2057 (19.6) |  |
| Great Britain | 1131 (10.8) |  |
| France | 770 (7.3) |  |
| China | 691 (6.6) |  |
| Netherland | 626 (6.0) |  |
| Australia | 586 (5.6) |  |
| Japan | 553 (5.3) |  |
| Germany | 401 (3.8) |  |
| Denmark | 337 (3.2) |  |
| Brazil | 324 (3.1) |  |
| other | 8904 (85.0) |  |

### Women authorship

From these authors, the gender API was able to propose a gender for 9596 authors. Among them, 3684 (34.1%) were women and 5912 (54.8%) were men with a median probability of 0.99 [0.97, 0.99], leaving 10.8% without defined gender (1159 because first name was not provided, 39 because the algorithm could not propose a gender). In the crude, non-imputed analysis (Supplementary Table 1), for the first author position there were 401 (36.8%) women and 546 (50%) men authors, with 13.2% of undetermined gender. For the last author position there were 284 (26.1%) women and 666 (61.2%) men authors, with 12.8% of undetermined gender.

Using GEE, the overall non adjusted estimated percentage of women authors across all positions and years was 39.8% (95%CI [38.4-41.2%]) overall, and 39.0% [37.5%-40.5%] if excluding pharmaceutical industry-affiliated authors.

In the adjusted analysis (table 3), compared to being a middle author, the odds ratio (OR) of being a woman author was lower for the last and penultimate position as author (0.72 [0.61-0.86]; corresponding to a -7.3 absolute percentage point (pp) decrease in woman author, and 0.70 [0.60-0.83]; -8.0pp respectively), and similar for first and second authorship.

**Table 3.**
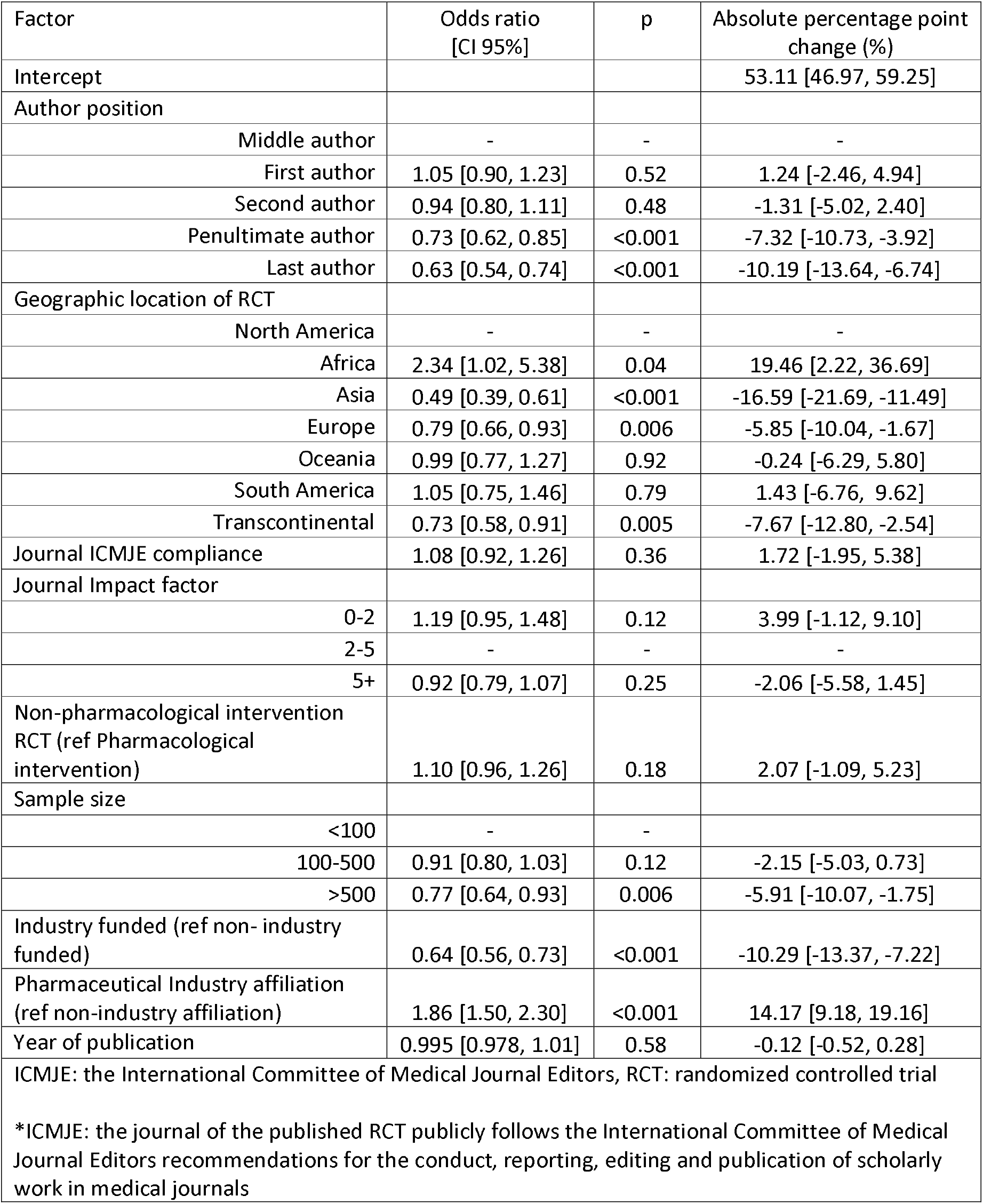
Factor associated with having a women as author in a randomized controlled trial (RCT)

Compared to RCT performed in North American, the OR of having a woman author was increased in African-based RCTs (OR 2.34 [95%CI 1.02-5.38]), + 19.5pp), and decreased in European based (0.79 [0.66, 0.93]; -5.9pp), transcontinental (0.73 [0.58, 0.91]; -7.7pp), and especially in Asian-based (0.49 [0.39-0.61]; -16.6pp) RCTs. There were also fewer women authors in industry-funded RCTs (OR 0.64 [0.56-0.73]; -10.3pp), and for RCTs of more than 500 patients (OR 0.77 [0.64, 0.93]; -5.9pp). Industry affiliation increased the chance of having a woman author (OR 1.88 [1.52-2.33]; +14.5pp). There were no changes in the proportion of women authors by year of publication.

Having a woman last-author increased the odds of having a woman author for the other authorship positions of 1.83 [1.46, 2.29], corresponding to an increase of woman authorship of 6.1pp [3.0-9.1].

## Discussion

The overall presence of women authors (including non-academic authors) was 39.5%. Several factors were associated with the presence or not of women authors.

First, when compared to North America, Europe, Asian and transcontinental RCTs had lower probability to have women authors, while African RCTS had a higher rate of women authorship. Industry-authors were also more likely to be women. While in North America the percentage of women in academia exceeds those of men (24), this is not the case in Europe (40), potentially explaining the lower odds of women in Europe being an author found in our analysis. On the other hand, the higher odds of women author in Africa is consistent with the higher rate of women rheumatologist (for example, more than 65% of rheumatologist are women in Egypt and Morocco (41,42), and represent more than 79% workforce in academic, with increasing trend in Arab countries (43)).

Second, RCTs sponsored by industry also had a lower representation of women authors. Industry-funded trials often involve high-profile studies, typically focusing on newly developed drugs that are critical to a company’s product pipeline. These trials generally require significant resources and offer substantial visibility in the medical community. The underrepresentation of women in these roles is particularly concerning given their importance and the potential for career advancement they represent.

Finally, positions such as last or penultimate author, which typically denote seniority and significant contributions to the research, were less likely to be occupied by women. This trend underscores the gender gap in senior roles within academic research. However, it’s important to note that while the first author position is generally considered to represent a more junior role, in the context of RCTs, the first author can often be a senior author as well. This variability in role significance highlights the complexity of interpreting author order and emphasizes the need for a thorough understanding when assessing gender disparities in research leadership. Our analysis also revealed that RCTs featuring a woman in the last author position were more likely to have a woman in the first author position as well. This pattern suggests a potential mentoring effect, where senior women researchers actively support and foster the development of emerging women scientists in their teams.

Contrary to findings from another study that evaluated gender trends in authorship across various types of rheumatology research (22), our analysis revealed no significant change in the trend of woman authorship in RCTs. The persistent stagnation in the representation of women year over year in RCTs despite the clear increase of women among rheumatologist worldwide highlights an urgent need for more effective strategies to close the gender gap in rheumatology research.

This research boasts several strengths. Our study utilizes a comprehensive database spanning almost 15 years. This extensive temporal scope allows us to observe and analyze trends over a significant period. We employed a solid methodological framework designed to address the challenges inherent in this type of research. This includes techniques for handling missing data and the probabilistic nature of gender determination. Such approaches enhance the reliability and validity of our findings, allowing for more precise assessments of gender disparities. Our study highlights the multifaceted dimensions of gender disparities in academic research and contributes valuable insights to the ongoing efforts aimed at achieving gender equity in scientific endeavors. By systematically documenting and analyzing these disparities, our work helps to identify key areas for intervention and supports the development of more inclusive research practices.

One limitation of this study is its reliance on binary gender data, which does not encompass non-binary or other gender identities. Additionally, the accuracy of gender determination from names, while robust, carries inherent limitations that could affect the interpretation of results.

The stagnation in gender parity within rheumatology RCTs authorship calls for a concerted effort to understand and dismantle the barriers to equity. Implementing structured policies and supporting women through mentorship and leadership opportunities are crucial steps towards a more inclusive and dynamic research environment. Such initiatives are not only essential for fostering gender equity but also for enhancing the quality of scientific research through diverse perspectives. The possible role model effect provides a compelling argument for initiatives aimed at promoting women into leadership positions within the academic and clinical research settings of rheumatology.

## Data Availability

All data produced in the present study are available upon reasonable request to the authors

## Notes

### Competing Interest Statement

The authors have declared no competing interest.

### Funding Statement

This study did not receive any funding

## References

1. Pell AN. Fixing the leaky pipeline: women scientists in academia. J Anim Sci [Internet]. 1 nov 1996 [cité 23 oct 2023];74(11):2843⍰8. Disponible sur: 10.2527/1996.74112843x

2. Spoon K, LaBerge N, Wapman KH, Zhang S, Morgan AC, Galesic M, et al. Gender and retention patterns among U.S. faculty. Sci Adv [Internet]. 20 oct 2023 [cité 23 oct 2023];9(42):eadi2205. Disponible sur: https://www.science.org/doi/10.1126/sciadv.adi2205

3. Hechtman LA, Moore NP, Schulkey CE, Miklos AC, Calcagno AM, Aragon R, et al. NIH funding longevity by gender. Proc Natl Acad Sci [Internet]. 31 juill 2018 [cité 23 oct 2023];115(31):7943⍰8. Disponible sur: https://www.pnas.org/doi/full/10.1073/pnas.1800615115

4. Larivière V, Ni C, Gingras Y, Cronin B, Sugimoto CR. Bibliometrics: Global gender disparities in science. Nature [Internet]. déc 2013 [cité 13 oct 2023];504(7479):211⍰3. Disponible sur: https://www.nature.com/articles/504211a

5. Ley TJ, Hamilton BH. The Gender Gap in NIH Grant Applications. Science [Internet]. 5 déc 2008 [cité 23 oct 2023];322(5907):1472⍰4. Disponible sur: https://www.science.org/doi/10.1126/science.1165878

6. Mayer SJ, Rathmann JMK. How does research productivity relate to gender? Analyzing gender differences for multiple publication dimensions. SCIENTOMETRICS [Internet]. déc 2018 [cité 13 oct 2023];117(3):1663⍰93. Disponible sur: https://www.webofscience.com/api/gateway?GWVersion=2&SrcAuth=DOISource&SrcApp=WOS&KeyAID=10.1007%2Fs11192-018-2933-1&DestApp=DOI&SrcAppSID=EUW1ED0F2DkfkQTxI7WBVTaTWEVbg&SrcJTitle=SCIENTOMETRICS&DestDOIRegistrantName=Springer-Verlag

7. Silver JK. Gender equity on journal editorial boards. The Lancet [Internet]. 18 mai 2019 [cité 13 oct 2023];393(10185):2037⍰8. Disponible sur: https://www.thelancet.com/journals/lancet/article/PIIS0140-6736(19)31042-6/fulltext#back-bib3

8. Bendels MHK, Müller R, Brueggmann D, Groneberg DA. Gender disparities in high-quality research revealed by Nature Index journals. PLOS ONE [Internet]. 2 janv 2018 [cité 23 oct 2023];13(1):e0189136. Disponible sur: https://journals.plos.org/plosone/article?id=10.1371/journal.pone.0189136

9. Ross MB, Glennon BM, Murciano-Goroff R, Berkes EG, Weinberg BA, Lane JI. Women are credited less in science than men. Nature [Internet]. août 2022 [cité 11 janv 2024];608(7921):135⍰45. Disponible sur: https://www.nature.com/articles/s41586-022-04966-w

10. Sebo P, Clair C. Gender gap in authorship: a study of 44,000 articles published in 100 high-impact general medical journals. Eur J Intern Med [Internet]. 1 mars 2022 [cité 23 oct 2023];97:103⍰5. Disponible sur: https://www.ejinme.com/article/S0953-6205(21)00313-7/fulltext

11. Madsen EB, Nielsen MW, Bjørnholm J, Jagsi R, Andersen JP. Author-level data confirm the widening gender gap in publishing rates during COVID-19. eLife. 16 mars 2022;11:e76559.

12. Gayet-Ageron A, Messaoud KB, Richards M, Schroter S. Female authorship of covid-19 research in manuscripts submitted to 11 biomedical journals: cross sectional study. BMJ [Internet]. 6 oct 2021 [cité 4 oct 2023];375:2288. Disponible sur: https://www.bmj.com/content/375/bmj.n2288

13. Hart KL, Perlis RH. Trends in Proportion of Women as Authors of Medical Journal Articles, 2008-2018. JAMA Intern Med [Internet]. 1 sept 2019 [cité 23 oct 2023];179(9):1285⍰7. Disponible sur: 10.1001/jamainternmed.2019.0907

14. West JD, Jacquet J, King MM, Correll SJ, Bergstrom CT. The Role of Gender in Scholarly Authorship. PLOS ONE [Internet]. 22 juill 2013 [cité 13 oct 2023];8(7):e66212. Disponible sur: https://journals.plos.org/plosone/article?id=10.1371/journal.pone.0066212

15. Mueller C, Wright R, Girod S. The publication gender gap in US academic surgery. BMC Surg. 14 févr 2017;17(1):16.

16. Schumacher C, Eliades T, Koletsi D. Gender gap in authorship within published orthodontic research. An observational study on evidence and time-trends over a decade. Eur J Orthod [Internet]. 1 oct 2021 [cité 13 oct 2023];43(5):534⍰43. Disponible sur: 10.1093/ejo/cjab036

17. Prunty M, Rhodes S, Sun H, Psutka SP, Mishra K, Kutikov A, et al. Redefining the Gender Gap in Urology Authorship: An 18-Year Publication Analysis. Eur Urol Focus [Internet]. 1 sept 2022 [cité 23 oct 2023];8(5):1512⍰9. Disponible sur: https://www.sciencedirect.com/science/article/pii/S2405456921003114

18. Silver JK, Poorman JA, Reilly JM, Spector ND, Goldstein R, Zafonte RD. Assessment of Women Physicians Among Authors of Perspective-Type Articles Published in High-Impact Pediatric Journals. JAMA Netw Open [Internet]. 20 juill 2018 [cité 13 oct 2023];1(3):e180802. Disponible sur: 10.1001/jamanetworkopen.2018.0802

19. Abraham RR, Adisa O, Owen ME, Iqbal F, Sulaiman K. Evaluation of gender trends in first authorship in nephrology publications in four major US journals in the last decade. J Nephrol. juin 2023;36(5):1395⍰400.

20. Purdy ME, Zmuda BN, Owens AM, Choudhary V, Olsen RC, Bader JO, et al. Gender differences in publication in emergency medicine journals. Am J Emerg Med [Internet]. 1 nov 2021 [cité 23 oct 2023];49:338⍰42. Disponible sur: https://www.sciencedirect.com/science/article/pii/S0735675721005234

21. Dalal NH, Chino F, Williamson H, Beasley GM, Salama AKS, Palta M. Mind the gap: Gendered publication trends in oncology. Cancer. 15 juin 2020;126(12):2859⍰65.

22. Levinsky Y, Vardi Y, Gafner M, Cohen N, Mimouni M, Scheuerman O, et al. Trend in women representation among authors of high rank rheumatology journals articles, 2002-2019. Rheumatol Oxf Engl. 3 nov 2021;60(11):5127⍰33.

23. Hassan N, Mens LJ van, Kiltz U, Andreoli L, Delgado-Beltran C, Ovseiko PV, et al. Gender equity in academic rheumatology: is there a gender gap at European rheumatology conferences? RMD Open [Internet]. 1 mars 2022 [cité 23 oct 2023];8(1):e002131. Disponible sur: https://rmdopen.bmj.com/content/8/1/e002131

24. Jorge A, Bolster M, Fu X, Blumenthal DM, Gross N, Blumenthal KG, et al. The Association Between Physician Gender and Career Advancement Among Academic Rheumatologists in the United States. Arthritis Rheumatol [Internet]. 2021 [cité 23 oct 2023];73(1):168⍰72. Disponible sur: https://onlinelibrary.wiley.com/doi/abs/10.1002/art.41492

25. Khursheed T, Ovseiko PV, Harifi G, Badsha H, Cheng YK, Hill CL, et al. Gender equity in rheumatology leadership in the Asia-Pacific. Rheumatol Adv Pract [Internet]. 6 sept 2022 [cité 3 juill 2024];6(3):rkac087. Disponible sur: 10.1093/rap/rkac087

26. Khursheed T, Harifi G, Ovseiko PV, Shekar HG, Badsha H, Gupta L. Is there a gender gap in global rheumatology leadership? Rheumatology [Internet]. 1 avr 2023 [cité 3 juill 2024];62(4):e107.#x2370;8. Disponible sur: 10.1093/rheumatology/keac499

27. Bagga E, Stewart S, Gamble GD, Hill J, Grey A, Dalbeth N. Representation of Women as Authors of Rheumatology Research Articles. ARTHRITIS Rheumatol [Internet]. janv 2021 [cité 13 oct 2023];73(1):162⍰7. Disponible sur: https://www.webofscience.com/api/gateway?GWVersion=2&SrcAuth=DOISource&SrcApp=WOS&KeyAID=10.1002%2Fart.41490&DestApp=DOI&SrcAppSID=EUW1ED0F2DkfkQTxI7WBVTaTWEV bg&SrcJTitle=ARTHRITIS+%26+RHEUMATOLOGY&DestDOIRegistrantName=Wiley+%28John+Wiley+%26+Sons%29

28. Hofstra B, Kulkarni VV, Munoz-Najar Galvez S, He B, Jurafsky D, McFarland DA. The Diversity-Innovation Paradox in Science. Proc Natl Acad Sci U S A. 28 avr 2020;117(17):9284⍰91.

29. Sebo P, Schwarz J, Achtari M, Clair C. Women Are Underrepresented Among Authors of Retracted Publications: Retrospective Study of 134 Medical Journals. J Med Internet Res. 6 oct 2023;25:e48529.

30. Mongin D, Buitrago-Garcia D, Capderou S, Agoritsas T, Gabay C, Courvoisier DS, et al. Prospective Registration of Trials: Where we are, why, and how we could get better [Internet]. medRxiv; 2024 [cité 21 août 2024]. p. 2024.07.24.24310935. Disponible sur: https://www.medrxiv.org/content/10.1101/2024.07.24.24310935v1

31. Cochrane. The Cochrane Highly Sensitive Search Strategies for identifying randomized trials in MEDLINE. In. Disponible sur: https://handbook-5-1.cochrane.org/chapter_6/6_4_11_1_the_cochrane_highly_sensitive_search_strategies_for.htm

32. Mongin D, Buitrago-Garcia D, Capderou S, Agoritsas T, Gabay C, Courvoisier DS, et al. Prospective Registration of Trials: Where we are, why, and how we could get better [Internet]. 2024 [cité 20 août 2024]. Disponible sur: http://medrxiv.org/lookup/doi/10.1101/2024.07.24.24310935

33. Santamaría L, Mihaljević H. Comparison and benchmark of name-to-gender inference services. PeerJ Comput Sci [Internet]. 16 juill 2018 [cité 4 oct 2023];4:e156. Disponible sur: https://peerj.com/articles/cs-156

34. Sebo P. Performance of gender detection tools: a comparative study of name-to-gender inference services. J Med Libr Assoc JMLA [Internet]. [cité 29 sept 2023];109(3):414⍰21. Disponible sur: https://www.ncbi.nlm.nih.gov/pmc/articles/PMC8485937/

35. Cheung YB. A modified least-squares regression approach to the estimation of risk difference. Am J Epidemiol. 1 déc 2007;166(11):1337⍰44.

36. Rubin DB. Multiple Imputation for Nonresponse in Surveys [Internet]. John Wiley & Sons, Ltd; 1987 [cité 19 juill 2021]. Disponible sur: https://onlinelibrary.wiley.com/doi/abs/10.1002/9780470316696.fmatter

37. R Core Team. R: A Language and Environment for Statistical Computing [Internet]. Vienna, Austria: R Foundation for Statistical Computing; 2019. Disponible sur: https://www.R-project.org

38. Højsgaard S, Halekoh U, Yan J. The R Package geepack for Generalized Estimating Equations. J Stat Softw [Internet]. 22 déc 2005 [cité 6 juill 2020];15(1):1⍰11. Disponible sur: https://www.jstatsoft.org/index.php/jss/article/view/v015i02

39. Buuren S van, Groothuis-Oudshoorn K. mice: Multivariate Imputation by Chained Equations in R. J Stat Softw [Internet]. 12 déc 2011 [cité 15 sept 2022];45:1⍰67. Disponible sur: 10.18637/jss.v045.i03

40. Ovseiko PV, Gossec L, Andreoli L, Kiltz U, Mens L van, Hassan N, et al. Gender equity in academic rheumatology, current status and potential for improvement: a cross-sectional study to inform an EULAR task force. RMD Open [Internet]. 1 août 2022 [cité 3 juill 2024];8(2):e002518. Disponible sur: https://rmdopen.bmj.com/content/8/2/e002518

41. Dey D, Paruk F, Mody GM, Kalla AA, Adebajo A, Akpabio A, et al. Women in rheumatology in Africa. Lancet Rheumatol [Internet]. 1 oct 2022 [cité 3 juill 2024];4(10):e657.#x2370;60. Disponible sur: https://www.thelancet.com/journals/lanrhe/article/PIIS2665-9913(22)00255-7/fulltext

42. Rheumatology TL. The road to gender equity must be paved with data. Lancet Rheumatol [Internet]. 1 oct 2022 [cité 3 juill 2024];4(10):e647. Disponible sur: https://www.thelancet.com/journals/lanrhe/article/PIIS2665-9913(22)00264-8/fulltext

43. Hmamouchi I, Ziadé N, Kibbi LE, Polyakov S, Arayssi T. Promising trends in authorship by Arab women in rheumatology. Lancet Rheumatol [Internet]. 1 oct 2022 [cité 3 juill 2024];4(10):e660.⍰2. Disponible sur: https://www.thelancet.com/journals/lanrhe/article/PIIS2665-9913(22)00220-X/fulltext

